# Germline Variants in Centromere Binding Protein 126 Predispose to Glioblastoma

**DOI:** 10.64898/2026.07.20.26358470

**Authors:** Paul Renauer, Bianca-Maria Marin, Adrian Melo-Carrillo, Mindy Carpenter, Linbin Zhang, MacLean Nasrallah, Jennifer Morrissette, Steven Brem, Yuan Rong, Alex M. Miller, Daniel A. Orringer, Matthew Bainbridge, Melissa Bondy, Sidi Chen, Roel G.W. Verhaak

## Abstract

**Background:** Glioblastoma (GBM) is the most common primary malignant brain tumor in adults. While only 5% of GBM cases arise in a familial context, the genetic basis of familial GBM remains unresolved in most affected clusters, suggesting that important susceptibility variants may reside outside recognized cancer-predisposition genes. Proband-based genomic studies of such families provide a rational path to discover rare inherited variants with large biological effects, prioritize candidate genes for functional validation, and define early mechanisms of gliomagenesis that may not be apparent from studies of sporadic tumors alone.

**Methods:** In this study, we investigated a family with GBM clustering spanning two generations. To investigate the possibility of shared germline susceptibility, we enrolled the proband and their two confirmed affected relatives in our study and performed whole-genome sequencing on available whole blood and tumor samples. Rare coding variants shared among the three study participants were identified and the genes harboring these variants were functionally interrogated with pooled loss-of-function CRISPR screens in human neural progenitor cells (NPCs) in vitro and in heterotopic xenograft models.

**Results:** Rare coding variants were identified in 139 candidate genes, and functional genetic screens of human NPCs in heterotopic xenograft models revealed Centrosomal Protein of 126 kDa (CEP126), as the top hit. Genetic disruption of CEP126 conferred a survival and tumorigenic advantage in neural progenitor cells.

**Conclusion:** Together, our results establish a functional framework for interrogating rare cancer germline variants and highlights CEP126 as a biologically tractable GBM predisposition gene for future mechanistic and genetic validation.

## INTRODUCTION

Glioblastoma (GBM) is the most common primary malignant brain tumor in adults, with a median age at diagnosis of 64 years^1^. Unlike colorectal, breast, lung, prostate or cervical cancers, there are currently no validated screening strategies for early GBM detection^2^. Most patients are diagnosed only after the onset of neurological symptoms, at which point the tumors are already clinically and molecularly complex^3^. Identifying heritable GBM predisposition factors could deepen our understanding of gliomagenesis and inform surveillance and early intervention strategies for high-risk individuals.

While having a first-degree relative with GBM confers a two-fold increased risk^4^, only approximately 5% of GBM cases arise in a familial context^5–7^. A minority of these familial cases (∼1%) develop within the context of hereditary cancer predisposition syndromes, including Li-Fraumeni (germline *TP53* mutations), Lynch syndrome (germline *MMR* mutations), familial adenomatous polyposis syndrome (germline *APC* mutations), and neurofibromatosis type 1 (germline *NF1* mutations)^8^. In these syndromes, GBMs typically occur alongside other malignancies, reflecting broad defects in genome maintenance and tumor suppressive pathways. However, other families show GBM clustering even in the absence of other cancer types, indicating the existence of GBM-specific susceptibility mechanisms. Genome-wide association studies (GWAS) have identified 27 loci associated with a modest increase in GBM risk^7,9–13^, and familial studies conducted by the Gliogene Consortium have identified additional predisposition genes^6,14–16^, including *POT1*, *HERC2*, *DMBT1*, and *HP1BP3*^6,14–16^. Despite these advances, the genetic basis of many familial GBM cases remains unresolved, underscoring the need for systematic approaches to identify, prioritize, and functionally interrogate rare germline variants.

Recent advances in CRISPR-Cas9 genome editing have provided powerful tools to interrogate the effects of candidate cancer genes^17–19^. In GBM research, CRISPR-based models have mostly focused on disrupting canonical tumor suppressors or activating established oncogenes in murine neural stem or progenitor cells^20–23^. Functional studies interrogating germline variants identified in GBM families have typically introduced the variants in combination with engineered loss of key GBM drivers, such as *TP53*, or *NF1*^14^. As a result, less is known about whether rare inherited variants can independently create early cellular advantages that precede acquisition of canonical GBM driver alterations.

In this study, we investigated a family with GBM clustering spanning two generations, suggestive of a hereditary predisposition. The proband, defined as the first affected individual to present for research evaluation, had been diagnosed with GBM in their early 40s, which is notably younger than the median age for sporadic GBM. Review of the proband’s family history revealed that a first degree relative (father) and a second-degree relative (paternal uncle) had also been diagnosed with GBM. Proband denied any family history of other malignancies. The occurrence of GBM across multiple generations, combined with the proband’s early age of onset, raised suspicion for a shared germline susceptibility among the affected relatives.

To investigate this possibility, we enrolled the proband and their two confirmed affected relatives in our study and performed whole-genome sequencing on available whole blood and tumor samples. Germline variants shared among the three study participants were identified, systematically filtered and functionally interrogated. Our results establish a functional framework for interrogating rare cancer germline variants and identify Centrosomal Protein of 126 kDa (*CEP126*), a microtubule-associated protein, as a candidate GBM predisposition gene. Using functional genetics, we demonstrate that the K605R germline mutation has a hypomorphic effect on function, and CEP126 loss-of-function confers a proliferative and tumorigenic advantage to neural progenitor cells (NPCs).

## MATERIALS AND METHODS

### Whole genome sequencing of patient samples

Peripheral blood DNA was extracted using the DNeasy Blood and Tissue kit (Qiagen) according to the manufacturer’s instructions. Tumor DNA was isolated from either frozen tumor tissue using QIAamp DNA mini kit (Qiagen) or from formalin-fixed paraffin-embedded (FFPE) tumor sections using the QIAamp DNA FFPE kit (Qiagen). Areas with the highest tumor content were selected for DNA extraction by a board-certified pathologist based on hematoxylin and eosin-stained section.

Library preparation and paired-end whole genome sequencing were performed at a mean sequencing depth was 30X for germline sample and 100X for tumor samples. Sequencing reads were aligned to human reference genome GRCh37/hg19 using BWA-MEM, and Picard and GATK (v4.1.0.0) were used to sort, mark duplicate reads and recalibrate base quality scores according to GATK Best Practices^24^.

### Genetic variant calling

Germline variants were identified using the local re-assembly *HaplotypeCaller* algorithm implemented in GATK (v4.1.0.0). For the proband, germline variants were identified from peripheral blood DNA. For the affected relatives, in absence of matched normal DNA sample, germline variants were inferred from tumor DNA. Joint genotyping was performed across all samples to generate a combined multi-sample VCF, enabling sensitive detection of shared variants and accurate genotype assignment. Somatic variants were identified from all three tumor samples using MuTect2 in tumor-only mode^25^.

### Genetic variant classification and annotation

VCF files containing germline variant calls from the three affected individuals were directly shared with AiLife Diagnostics for independent clinical evaluation. Variant classification was performed using AiLife Diagnostics’ in-house developed pipeline in accordance with the American College of Medical Genetics and Genomics (ACMG) guidelines^26^. A curated list of glioma susceptibility loci, together with their reported risk allele frequencies and corresponding effect sizes (odds ratios) related to GBM development, was obtained from published GWAS study data^13,27^. Shared germline variants identified in the study family were intersected with these glioma-associated risk loci to determine overlap. Germline variants shared across all three affected individuals were annotated using Ensembl Variant Predictor (Release 115)^28^. Annotations included genomic, gene assignment based on RefSeq transcripts, predicted functional consequence, and population allele frequencies derived from the Genome Aggregation Database (gnomAD v4)^29^.

### Filtering for novel glioma susceptibility genes

Shared germline variants were filtered using a rare disease-oriented approach to prioritize candidate susceptibility genes with high penetrance. Variants were first filtered based on population frequency to retain only novel variants not reported in gnomAD and variants present at a minor allele frequency (MAF) < 0.01 across all reported populations, using the “max_AF” allele frequency metric. Variants were subsequently filtered based on location to maintain only those in coding exons based on RefSeq annotation. Finally, synonymous variants predicted not to alter protein structure or canonical splice sites were excluded.

### Functional annotation of putative glioma susceptibility genes

Gene Ontology Biological Process overrepresentation analysis was performed in R using clusterProfiler (v4.10.1) to assess the enrichment of biological processes among the 139 candidate genes relative to the background of all protein-coding genes. P values were adjusted for multiple hypothesis testing using the Benjamini-Hochberg method. No pathways met significance at an adjusted p value < 0.05. For visualization purposes, genes were manually grouped into broad biological themes based on their predicted cellular functions.

### Cell lines and culture

Human neural progenitor cells (StemCell Technologies) were cultured in StemDIFF Neural Progenitor Medium (StemCell Technologies) on matrigel-coated plates (Corning). Human embryonic kidney (HEK) 293T cells were cultured in high glucose DMEM (Gibco) supplemented with 10% fetal bovine serum (Avantor) and 1% penicillin/streptomycin (Gibco). All cells were cultured in a humidified incubator at 37°C with 5% CO_2_. Cells were routinely tested and were negative for mycoplasma contamination (MycoAlert Mycoplasma Detection Kit, Lonza).

### sgRNA library design

A custom CRISPR-Cas9 sgRNA library targeting the 139 candidate genes was designed using the Genetic Perturbation Platform (Broad Institute). Six sgRNAs per gene were selected based on unscaled RuleSet3 on-target scores > 0. sgRNAs were prioritized based on transcript-based proximity to the identified germline variant(s), with the requirement that at least 3 sgRNAs per gene had predicted cutting efficiency > 20%. Negative selection controls included sgRNAs targeting 10 essential genes (*RPA3, POLR2B, PCNA, DBR1, PLK1, RPL3, GAPDH, KIF11, PSMB1, EEF2*), while positive selection control sgRNAs targeted tumor suppressor genes (*TP53, PTEN, NF1, CDKN2A, RB1*). The library additionally included 200 intergenic regions-targeting sgRNAs as Cas9 cutting controls, and 200 non-targeting sgRNAs as neutral controls. CRISPR spacer sequences were synthesized by Twist Bioscience.

### Library cloning and lentiviral packaging

Library oligonucleotides were cloned into the pSC007 lentiviral vector using Gibson assembly (New England Biolabs). Library lentiviral particles were produced in HEK293T cells by co-transfection of either the Cas9-expressing vector (pHK014) or the sgRNA library vector (pSC007) with pPAX2 and pMD2.G packaging plasmids using LipoD 293T transfection reagent (Signagen). Plasmids were generously provided by the Chen laboratory.

Viral supernatant was collected at 48 h and 72 h post-transfection and concentrated using a 4x Lenti-Concentrator following the MD Anderson Functional Genomics Core protocol. Concentrated virus was resuspended in DMEM/F12 and flash-frozen in liquid nitrogen.

### CRISPR-Cas9 and sgRNA library transduction in primary NPCs

Primary NPCs were plated on matrigel-coated 6-well plates (Corning) and allowed to recover overnight. NPCs were first transduced with Cas9 lentivirus at a multiplicity of infection (MOI) of 0.8 for 24 h, followed by media replacement. After an additional 24 h, cells were transduced with pooled sgRNA library lentivirus at an MOI of 0.2 for 24 h. Following a 24 h recovery period, Cas9- and sgRNA library-expressing cells were selected using 1 μg/ml puromycin (InvivoGen) and 5 μg/ml blasticidin (InvivoGen) for 3 days with daily media replacement. Surviving cells were either used for the CRISPR screens or cryopreserved as pre-screen baseline samples.

### Short term survival assay

NPC-Cas9 cells expressing either the sgRNA library (NPC-Cas9-Library) or a neutral *AAVS1*-targeting sgRNA (NPC-Cas9-*AAVS1*) were dissociated and plated into white matrigel-coated 96-well plates (Thermo Fisher) and cultured under standard conditions. Cell proliferation was measured after 72 h with CellTiter-Glo 3D Cell Viability Assay (Promega) according to manufacturer instructions and luminescence was measured using a Tecan Infinite 200 PRO plate reader.

### Functional genomic screens

For in vitro screening, library-transduced NPCs (NPC-Cas9-Library) were cultured for 14 days under standard growth conditions prior to genomic DNA extraction. For in vivo CRISPR screens, 4 × 10^6^ NPC-Cas9-Library cells were mixed 1:1 with matrigel (Corning) and injected subcutaneously into the flanks of 12-14-week-old female Nu/Nu nude mice (n=5; Charles River). Control mice (n=5) were injected with NPC-Cas9-*AAVS1* cells. Tumor formation was monitored for 45 days post-injection via caliper measurements. At endpoint, tumors were harvested, mechanically dissociated, and digested in 3 mL collagenase IV (Gibco) for 60 minutes at 37°C prior to DNA extraction. All animal experiments were approved by the Yale University Institutional Animal Care and Use Committee.

### Library sequencing

Genomic DNA was isolated using the DNeasy Blood and Tissue kit (Qiagen) following the manufacturer’s protocol. sgRNA sequences were amplified using a two-step nested PCR strategy. In the first PCR, sgRNA cassette region was amplified with Phusion Flash High Fidelity Master Mix (Thermo Scientific). The product was subsequently amplified in the second PCR using barcoded adapter primers for sequencing. PCR products were gel-purified from a 2% agarose gel using the QIAquick Gel Extraction Kit (Qiagen). Paired-end sequencing of pooled sgRNA libraries was performed by the Yale Center for Genomic Analysis with a NovaSeq X (Illumina).

### CRISPR screen data analysis

Adapter sequences were trimmed from raw FASTQ files via Cutadapt v3.41 with a 10% error tolerance and resulting reads shorter than 15 nucleotides were discarded. The remaining reads were aligned to a reference containing all sgRNA spacer sequences using Bowtie (v1.3.02) with parameters -v0, -m1 --best. sgRNA counts were processed and analyzed using the SAMBA R package (v1.3.0). Positive selection scores were calculated to identify genes whose knockout promoted cell growth and/or survival. Statistical significance was assessed using the false discovery rate (FDR), with significant hits defined as FDR < 0.05.

### Single-nucleus RNA sequencing analysis

Previously reported processed data from primary untreated GBMs (n=52) were analyzed (Gene Expression Omnibus: accession GSE274546)^30^. Gene expression values were log-normalized and scaled using *NormalizeData* and *ScaleData* Seurat (v.4.0.4) functions, respectively. Cell identities and states were assigned as previously annotated^30,31^. Malignant nuclei enriched for the cilia-like metaprogram were subset using Seurat. Gene-wise Pearson correlation coefficients were calculated between expression of CEP126 and all other genes detected in the cilia-like malignant nuclei using log-normalized RNA counts from Seurat. P values were computed from the Pearson correlation statistic and adjusted using the Benjamini-Hochberg method. Genes with FDR < 0.05 were considered significant and ranked by correlation coefficient.

### In silico pathogenicity prediction

The predicted functional impact of the CEP126^K605R^ variant was assessed using AlphaMissense^32^. The variant was queried against the UniProt reference sequence for human CEP126, and the reported pathogenicity score (range 0-1) was used to classify the variant as benign, uncertain, or likely pathogenic according to the tool’s predefined thresholds.

### Regulatory effects prediction

Potential regulatory consequences of the CEP126^K605R^ variant were assessed using AlphaGenome^33^. The variant was assessed for predicted changes in gene expression, chromatic accessibility, as well as overlap with known regulatory regions based on Ensembl regulatory build.

### Immunofluorescence Microscopy

Immunofluorescence staining was performed on NPCs cultured on chamber slides (8-well, Soda Lime Glass) (Nunc Lab-Tek). Cells were washed with phosphate-buffered saline (PBS) and fixed in 4% paraformaldehyde for 15 min at room temperature. After fixation, cells were washed with PBS and permeabilized with 0.3% Triton X-100 in PBS for 20 min. Samples were then blocked in PBS containing 0.1% bovine serum albumin for 60 min at room temperature. Primary antibodies against human α-tubulin, γ-tubulin, and CEP126 proteins were diluted in blocking buffer and incubated overnight at 4°C. Cells were washed three times with PBS and incubated with fluorophore-conjugated secondary antibodies for 3 h at room temperature. DNA was counterstained with Hoechst 33342 (1:1000) for 10 min, followed by three PBS washes. Coverslips were mounted using ProLong™ Glass Antifade Mountant (Invitrogen) and imaged using a Stellaris 8 Falcon Confocal Microscope (Leica).

## RESULTS

### Identification of shared somatic and germline variants

To identify the genetic basis of GBM predisposition in our study cohort, we performed whole genome-sequencing on tumor tissue from the proband, a first degree relative (father), and a second-degree relative (paternal uncle), along with peripheral blood from the proband (**Figure 1B**). This design enabled us to identify both somatic alterations driving tumorigenesis and germline variants underlying inherited susceptibility.

**Figure 1.**
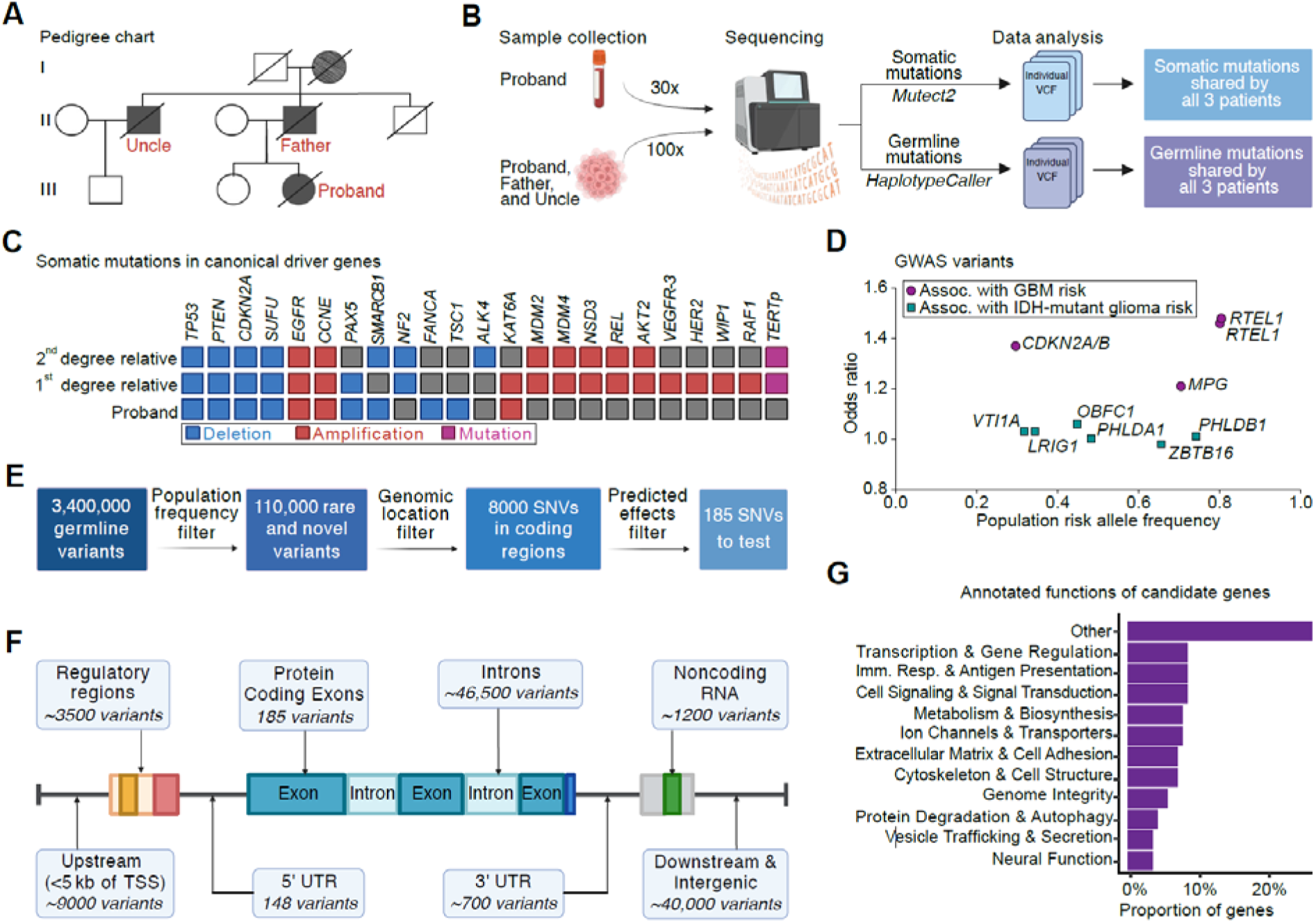
Analysis of shared somatic and germline variants. **A,** Pedigree chart of the GBM study family. Squares represent males and circles represent females. Solid shading designates individuals with clinically confirmed GBM, while diagonal hatching indicates suspected primary brain tumor. Deceased individuals are marked with a slash through the symbol. Family members enrolled in the study are indicated in red. **B,** Schematic for the identification of shared somatic and germline variants. **C,** Somatic alterations in canonical GBM- and cancer-associated driver genes detected in the three study participants. Blue boxes denote deletions, red boxes denote amplifications, purple indicates TERT promoter mutation, and grey indicates no detected alteration. **D,** Summary of the 10 GWAS risk variants shared by the study participants. Purple and green denote variants associated with risk for GBM and *IDH*- mutant gliomas, respectively. Variants are labeled with location and gene name, which was chosen based on the variant’s proximity to the gene or their hypothesized relationship^13^. Risk allele frequency was based on European samples for 1000 genomes project^9,29^. **E,** Schematic of the stepwise filtering strategy applied to germline variants shared across the three affected individuals. **F,** Genomic region distribution for rare and novel variants after filtering for population frequency and removing synonymous mutations from protein-coding exons. **G,** Bar plot of the functional classification of candidate GBM predisposition genes across major biological categories.

First, we analyzed somatic mutations to confirm tumor identity and the presence of molecular GBM features. Somatic alterations shared between the three study participants revealed canonical GBM alterations^34^, including deletion of *TP53*, *PTEN*, *CDKN2A*, and amplification of *EGFR*, consistent with their clinical GBM diagnosis (**Figure 1C**).

Next, we sought to identify the germline variants shared among the three affected relatives, who shared an Ashkenazi-Jewish ancestry (**Figure 1B**). For the proband, germline variants were called directly from blood DNA and tumor. Due to the absence of blood samples from the affected relatives, putative germline variants were called from their tumor samples. To minimize inclusion of somatic variants, analyses were restricted to variants shared across the three individuals and present at allele fractions consistent with constitutional germline variants^35^. In line with expected burden of human germline variation^29^, we identified approximately 3.4 million shared germline variants across the three study participants relative to the hg19 human reference genome. The common variants included ∼2,500,000 single nucleotide variants (SNV) and ∼900,000 insertions and deletions (indels). These variants were broadly distributed across the genome and served as the starting point for downstream filtering to identify candidate GBM predisposition genes.

### Evaluation of variants in cancer predisposition genes and GBM GWAS loci

After identifying shared germline variants, we asked whether they included any deleterious germline variants in known cancer-predisposition genes. A clinical hereditary cancer analysis performed following American College of Medical Genetics and Genomics (ACMG) guidelines^26^ (AiLife Diagnostics) identified no pathogenic or likely pathogenic variants in established cancer predisposition. There was a single variant of unknown significance (VUS) in the 3’ untranslated region of *MET* (c*3051C>T) shared by the three affected individuals. However, since oncogenic *MET* alterations are typically coding mutations associated with papillary renal carcinoma type I^36^, the detected VUS is unlikely to represent the causal driver of disease in the study family.

Next, we evaluated the presence of mutations in any glioma susceptibility loci identified in GWAS studies^7,9–13^. The three affected individuals shared ten GWAS variants, including 4 GWAS variants associated with an increased risk of developing GBM (*CDKN2A*/B, *MPG*, *RTEL1*) and 6 variants associated with *IDH*-mutant glioma (*LRIG1*, *OBFC1*, *PHLDA1*, *PHLDB1*, *VTI1A*, *ZBTB16*)^13^ (**Figure 1D**). However, all identified variants were common in European-ancestry populations (allele frequencies ∼0.3-0.8) and exhibited modest effect sizes related to GBM development risk (odds ratios < 1.5)^9^.

Together, these results suggest that the genetic basis of GBM predisposition in our study cohort is underlined by a variant in a novel gene, rather than a known cancer-associated gene or a glioma-associated GWAS risk locus.

### Nomination of novel GBM predisposition genes

In the absence of pathogenic variants in established cancer genes, we set out to identify novel candidate genes that may underlie glioma predisposition in our study family. To this end, we annotated and systematically filtered the 3.4 million shared variants based on genomic location, population frequency, and predicted molecular consequences (**Figure 1E**).

Given that glioma has an incidence of only 6 per 100,000 individuals^37^, we reasoned that the predisposing variant would be either novel or extremely rare in the general population. Therefore, we first filtered variants based on general population frequency, retaining only novel variants, not previously reported in the Genome Aggregation Database^38^ (gnomAD), and rare variants, present in less than 1% of individuals across any population listed in gnomAD. This resulted in a set of approximately 110,000 variants distributed across the genome (**Figure 1F**).

Since most disease-causing mutations are found in protein-coding regions^39^, we focused on the shared variants located within coding exons. We then excluded approximately 8,000 synonymous variants that were unlikely to affect protein structure and function. This stepwise filtering strategy resulted in a set of 185 potentially protein-altering SNVs (182 missense and 3 nonsense) across 139 genes (**Figure 1F**). No indel variants in coding regions passed the population filtering step.

Functional annotation of the 139 candidate genes revealed involvement in diverse biological processes (**Figure 1G**), including genome stability, transcriptional regulation, cellular metabolism, intracellular transport, cytoskeleton organization, and immune system signaling. However, no pathways were significantly enriched after correcting for multiple testing, reflecting the functional heterogeneity of the candidate set. Importantly, the candidate list included several genes previously implicated in tumor suppressor functions, including *DOCK2*^40^, *HERC1*^41^, *MGA*^42,43^, *PTPRS*^44^, and *RIF111*^45^, as well as genes reported to promote tumor progression, such as *ATG4C*^46^ or *WNK1*^47^.

### Functional CRISPR screening of candidate genes

Given the functional heterogeneity of the 139 candidate genes, we aimed to systematically interrogate the gene set in an unbiased manner. Since loss-of-function mutations in tumor suppressor genes underlie most cancer predisposition syndromes^48^, we hypothesized that the loss of one or more of these candidates would confer a selective growth or early oncogenic advantage to NPCs. To test this hypothesis, we generated a pooled CRISPR-Cas9 loss-of-function sgRNA library targeting all 139 genes. Each gene was represented by 6 sgRNAs designed to induce frameshift mutations proximal to the identified germline variant(s). The library incorporated additional control guides, including sgRNAs targeting canonical tumor suppressors (positive controls (*PTEN*, *TP53*, and *CDKN2A*)), sgRNAs targeting oncogenes and essential genes (negative controls), intergenic-targeting sgRNAs to control for Cas9 cutting effects, and non-targeting sgRNAs with minimal homology to the human genome.

In line with recent studies using NPCs to study gliomagenesis drivers^49,50^, we selected primary human NPCs as a biologically relevant background for the screen. NPCs were transduced with lentiviruses encoding Cas9 and the pooled sgRNA library at low multiplicity of infection to ensure predominantly single guide integration per cell (**Figure 2A**). To evaluate selective pressure in vitro, library-transduced NPCs were cultured under standard conditions for 14 days. Short-term cell viability assays revealed that library-transduced NPCs had significantly increased proliferation at day 5 compared to control NPCs expressing Cas9 and a neutral *AAVS1*-targeting sgRNA (**Figure 2B**), suggesting that disruption of one or more genes conferred an early growth advantage.

**Figure 2.**
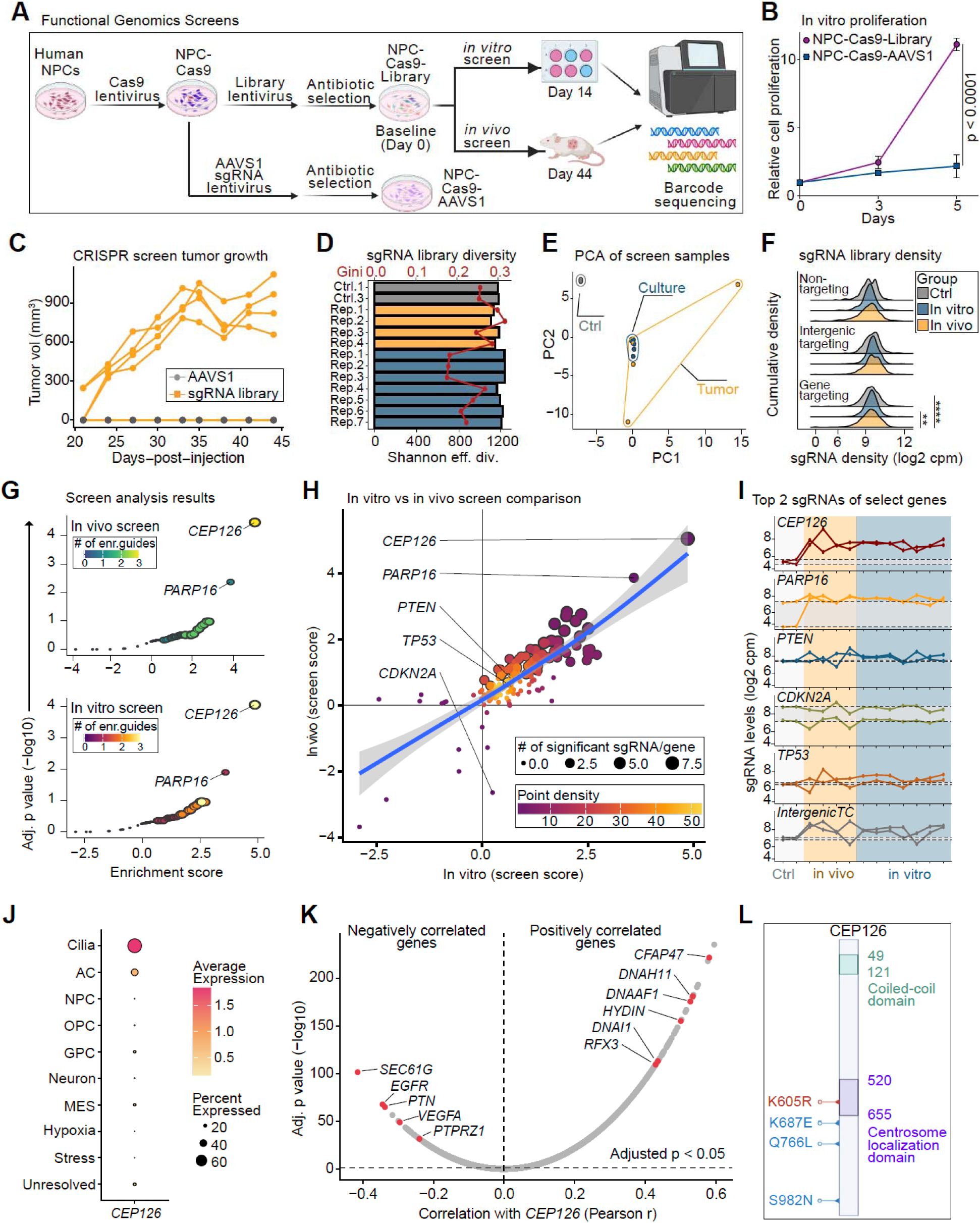
Functional genetics investigation identifies CEP126 as top tumorigenesis candidate. **A,** Schematic overview of in vitro and in vivo loss-of-function CRISPR screens. **B,** Line plots for proliferation of NPC-Cas9-Library and NPC-Cas9-AAVS1 cells, measured by CellTiter-Glo. Proliferation rates were normalized to day 1 values (n=3 biological replicates/condition). **C,** Tumor growth curves following heterotopic flank implantation of NPC-Cas9-Library and NPC- Cas9-AAVS1 cells into Nu/Nu mice. **D,** Bar plot of sgRNA library diversity across screen and control (baseline) samples, shown by Shannon effective diversity and Gini index metrics (bar and line, respectively). **E,** Plot of first two components of the principal components analysis (PCA) of samples, based on sgRNA values. **F,** Density distributions of sgRNA abundance for gene-targeting, non-targeting, and intergenic guides across baseline control and screen samples. The x-axis shows normalized sgRNA abundance (log2 counts per million; CPM). **G,** Volcano-style plot of gene-level enrichment scores from the (**top**) in vivo and (**bottom**) in vitro screens. Genes are plotted with respect to SAMBA enrichment scores and significance (FDR-adjusted p values). **H,** Plot comparing gene-level enrichment scores of in vitro and in vivo CRISPR screens. The blue line depicts a trendline fitted with a generalized additive model, and the 95% CI is shown in gray. **I,** Line plots of individual sgRNA levels for select genes across samples. The log2-CPM values for the top two sgRNAs (assessed from the screens) are shown for each depicted gene. For baseline comparisons, a gray horizontal rectangle presents the control sample values for each gene, and intergenic-targeting sgRNAs (95^th^ percentile) are shown at the bottom. **J,** Dot plot showing CEP126 expression across malignant cellular states in single-nucleus RNA sequencing data from primary untreated sporadic GBMs (n=52). Dot size represents the percentage of nuclei expressing CEP126 in each state, and color represents average expression. **K,** Gene-wise Pearson correlation analysis between CEP126 expression and all other genes within malignant cilia-like nuclei. Horizontal dashed line marks adjusted p value < 0.05, and vertical dashed line represents correlation r = 0. Positively correlated genes (**right**) included multiple canonical ciliogenesis and axonemal components. Negatively correlated genes (**left**) include canonical GBM-associated signaling genes. **L,** Schematic representation of CEP126 protein domains and location of identified germline variants. Red mutation denotes the variant identified in our study family, while blue variants are mutations found in families from the Gliogene consortium. Statistics were performed using parametric two-tailed t-test (**B**, at day 5), two-way ANOVA with Tukey’s post-hoc analysis (**F**), SAMBA screen analysis (**G-H**), and Pearson correlation analysis (**K**). Significance levels for panels F-H and K are presented with FDR-adjusted P values.

To assess acquisition of tumorigenic competence in vivo, library-transduced NPCs were heterotopically implanted into immunodeficient nude mice. Notably, pooled library-transduced NPCs formed tumors in 4 out of the 5 injected mice within 45 days. In contrast, NPCs-Cas9- AAVS1 cells failed to form tumors, indicating that disruption of one or more library-targeted genes was sufficient to enable malignant outgrowth in this context (**Figure 2C**).

To quantify changes in sgRNA representation across experimental conditions, sgRNA abundance was quantified by short-read sequencing at baseline (day 0), in vitro (day 14), and in vivo (day 45) samples. Screen-level QC showed adequate maintenance of sgRNA library diversity across replicates, with increased abundance skewing in selected samples (**Figure 2D**). Principal component analysis separated control, culture, and tumor samples, while guide-level density distributions demonstrated screen-specific shifts in gene-targeting but not control sgRNAs (**Figure 2E-F**), together indicating reproducible selection of sgRNA perturbations during in vitro and in vivo screening.

Screen data analysis by SAMBA^51^ identified two candidates with enrichment significantly across background. The most enriched candidate was *CEP126* (*KIAA1377*), both in vitro and in vivo (FDR-adjusted p = 8.84 × 10^−5^ and 3.6 × 10^−5^, respectively) (**Figure 2G-H**). Multiple *CEP126*- targeting sgRNAs were independently enriched across both biological contexts, supporting a gene-specific effect rather than an sgRNA-specific artifact (**Figure 2I**). A secondary candidate enriched both in vitro and in vivo was Poly(ADP-Ribose) Polymerase Family Member 16 (*PARP16*) (**Figure 2G-I**). However, guide-level analysis revealed that the effect was primarily driven by a single sgRNA, while another *PARP16*-targeting sgRNA exhibited significant depletion. This bidirectional effect of individual sgRNAs raises the possibility of guide-specific off-target effects or heterozygous editing outcomes and weakens the confidence in *PARP16* having tumor suppressor-like effects. Other candidate genes exhibited limited selective pressure upon deletion.

Collectively, these results identify *CEP126* the most robustly selected candidate in both CRISPR screens. The consistent multi-guide enrichment observed in vitro and in vivo nominates *CEP126* as a putative GBM predisposition gene whose loss confers a selective growth advantage and tumorigenic potential in human NPCs.

### Computational evaluation of CEP126^K605R^ variant

CEP126 has been previously characterized as a centrosomal protein required for primary cilium formation^52^, but its role in cancer remains undefined. To investigate the biological context of *CEP126* in GBM, we analyzed its expression in malignant nuclei from published single-nucleus RNA sequencing data of primary untreated GBMs^30,31,53^. Consistent with its established role in ciliogenesis, *CEP126* expression was highest in malignant cells expressing a cilia-like transcriptional metaprogram^30,31^ (**Figure 2J**). Furthermore, gene-wise correlation analysis within the cilia-like malignant cell population revealed that *CEP126* expression positively correlated with core components of the ciliary machinery, including multiple genes of the *DNAH* and *CFAP* families (**Figure 2K**). In contrast, *CEP126* expression was inversely correlated with several genes associated with oncogenic signaling, including *EGFR* (r = -0.344, FDR-adjusted p = 2.20 × 10^−68^) and *SEC61G* (r = -0.415, FDR-adjusted p = 3.47 × 10^−102^), which are co-amplified on chromosome 7 in ∼50% of GBM cases^54^. Negatively correlated genes also included *PTPRZ1* (r = -0.240, FDR-adjusted p = 2.63 × 10^−32^) and its ligand *PTN* (r = -0.337, FDR-adjusted p = 1.23 × 10^−65^), both of which are enriched in glioma stem-like cells^55^, as well as *VEGFA* (correlation = -0.295, FDR-adjusted p = 1.17 × 10^−49^), a key driver of GBM angiogenesis^56^.

We next examined the specific *CEP126^K^*^605^*^R^* variant shared by the three study participants. GnomAD analysis^57^ revealed the variant was rare in the general population, with a maximum allele frequency of 0.2%. To evaluate the variant’s potential functional impact, we queried AlphaMissense, a deep learning-based model that predicts the structural consequences of missense subsitutions^32^. K605R received an AlphaMissense substitution score of 0.096, suggesting the variant is unlikely to cause major structural effects. However, since the K605R variant resides within the centrosome-localization region of *CEP126* (residues 520-655)^52^, this prediction cannot exclude the possibility that the variant may exert other functional effects not captured by the structural prediction. We also investigated potential effects on regulatory elements using the AlphaGenome sequence-based model^33^. Consistent with its location in a protein-coding exon, AlphaGenome predicted no significant impact of the K605R variant on gene expression (log-fold change = -2 × 10^−5^) or chromatin accessibility (mean-effect = 4.4 × 10^−5^).

Lastly, we investigated *CEP126* variation within the Gliogene consortium dataset, which contains whole exome sequencing data from multiple cases of familial GBM^14^. Although no families harbored the K605R variant, three out of 189 independent families carried other rare nonsynonymous *CEP126* variants, including K687E, Q766L, and S982N (**Figure 2L**), indicating that rare *CEP126* variation is present across multiple familial GBM cases.

### CEP126 disruption enhances NPC survival and tumorigenesis

To determine whether *CEP126* disruption was sufficient to alter growth behavior, we generated Cas9-expressing primary NPCs and compared the phenotype in cells treated with *AAVS1* vs *CEP126*-targeting sgRNAs (top 2 sgRNAs from the screen, herein referenced as *CEP126*_sg2 and *CEP126*_sg5). Flow cytometry analyses demonstrated efficient loss of CEP126 protein expression from both *CEP126* sgRNAs (**Figure 3A**). We then assessed proliferation dynamics using a CellTrace Dilution Assay, which demonstrated that *CEP126* loss from either sgRNA significantly altered the cell cycle dynamics in low and intermediate but not highly proliferating cells (**Figure 3B**). This effect was accompanied by reduced KI-67 staining in *CEP126*-sgRNA populations (**Figure 3C**), suggesting that *CEP126* loss changes cell-cycle state rather than simply increasing the fraction of actively cycling cells at a single time point. To determine if this *CEP126*-mediated increase in slower proliferating cells was due to enhanced survival, we quantified apoptosis in the NPCs using Annexin-V and showed *CEP126* loss significantly decrease apoptosis relative to *AAVS1*-sgRNA controls (**Figure 3D**).

**Figure 3.**
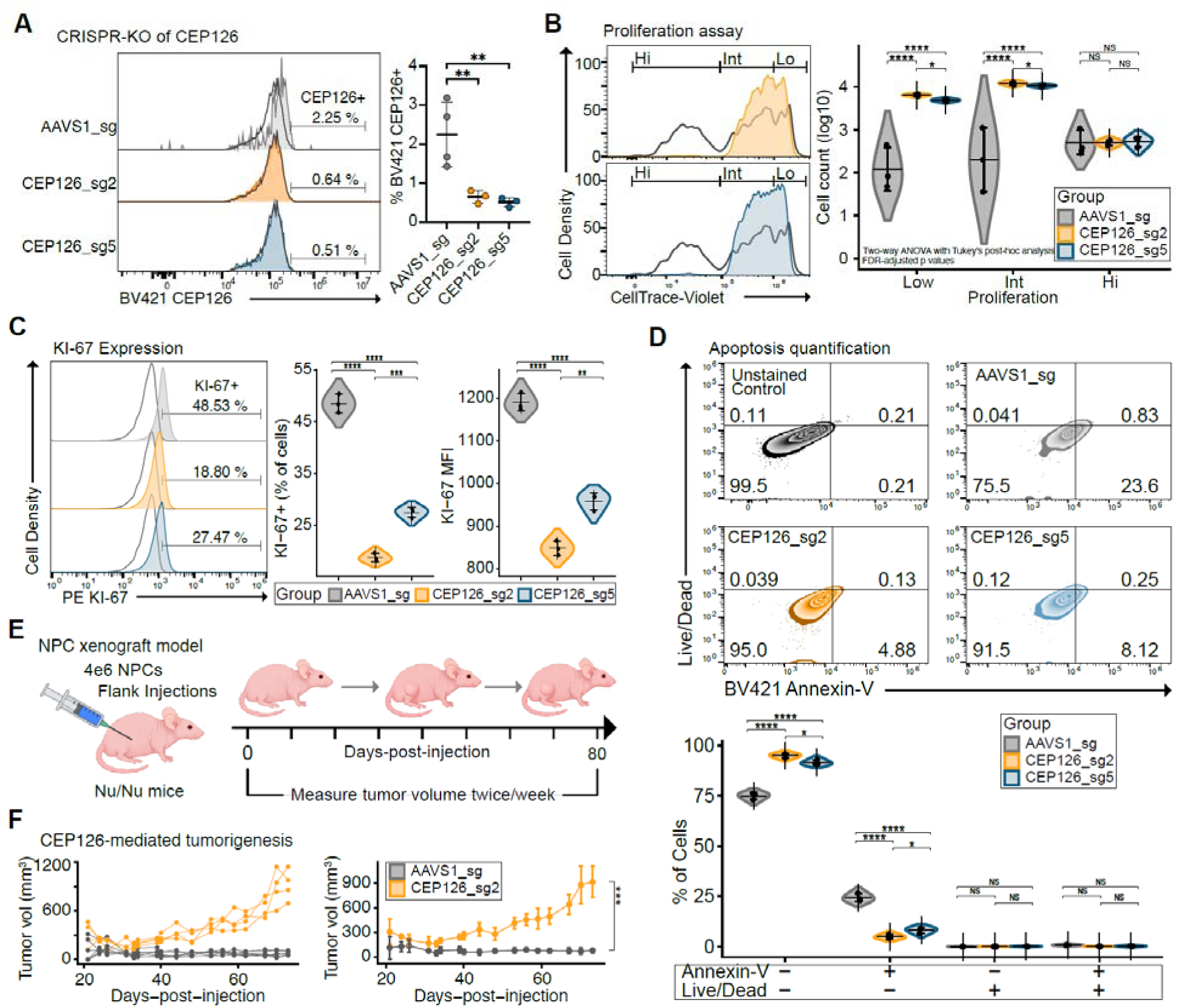
CEP126 disruption enhances NPC survival and tumorigenesis. **A,** Flow cytometry histogram quantifying the CEP126 protein levels in Cas9-expressing NPCs treated with sgRNAs targeting *AAVS1* (control) or the *CEP126* locus. **B,** Flow cytometry histogram of CellTrace proliferation assay. Low, intermediate, and highly proliferating cells are distinguished by CellTrace dye dilution across AAVS1, and CEP126 sgRNA-treated NPCs. **C,** Histograms of KI-67 protein expression across treatment groups. The violin plots on the right compare KI-67 positive cell percentages and geometric mean fluorescence intensities (MFIs). **D,** Cytometry plots showing apoptosis quantification (Annexin-V+Live/Dead- cells). **E,** Schematic of heterotropic xenograft of AAVS1 or CEP126 sgRNA NPCs into Nu/Nu mice. **F,** Individual and grouped tumor growth curves for the xenograft models. Statistics included one-way (**A** & **C**) and two-way (**B**, **D**, and **F**) ANOVA tests with Tukey’s post-hoc analyses, and statistical significance is presented by FDR-adjusted p values. Panels A-D included n=3 biological replicates, while panel F depicts n=4 CEP126 and n=6 AAVS1 sgRNA xenografted mice.

We next tested whether this growth advantage extended in vivo using a heterotopic xenograft model. Control and *CEP126*-targeted NPCs were implanted subcutaneously, and tumor volume was measured longitudinally (**Figure 3E**). In contrast to *AAVS1* sgRNA cells, the *CEP126*_sg2 cells formed tumors with increasing growth trajectories in all xenograft mice (**Figure 3F**). Together, these data indicate that *CEP126* disruption is sufficient to confer a reproducible growth and tumorigenic advantage in human NPCs.

### CEP126 loss alters centrosome-associated structures of neural progenitor cells

CEP126 regulates centrosome-associated microtubule organization, in part, through effects on pericentriolar satellite trafficking and γ-tubulin-associated centrosomal structures^58^. Since γ- tubulin is central to mitotic microtubule nucleation and proper spindle function^59^, we hypothesized that *CEP126* disruption may impair the centrosome-dependent processes required for faithful chromosome segregation.

To define the cellular consequences of *CEP126* disruption, we examined centrosome- and microtubule-associated phenotypes in *AAVS1* (control) vs *CEP126* sgRNA NPCs by immunofluorescence. In the control cells, CEP126 was detected as discrete puncta that localized near γ-tubulin-positive centrosomal structures, consistent with the expected centrosomal localization of CEP126 (**Figure 4A**). In contrast, *CEP126*-sgRNA cells showed marked reduction of CEP126 signal, including the puncta number, area, and mean intensity, confirming efficient perturbation at the protein level (**Figure 4B**).

**Figure 4.**
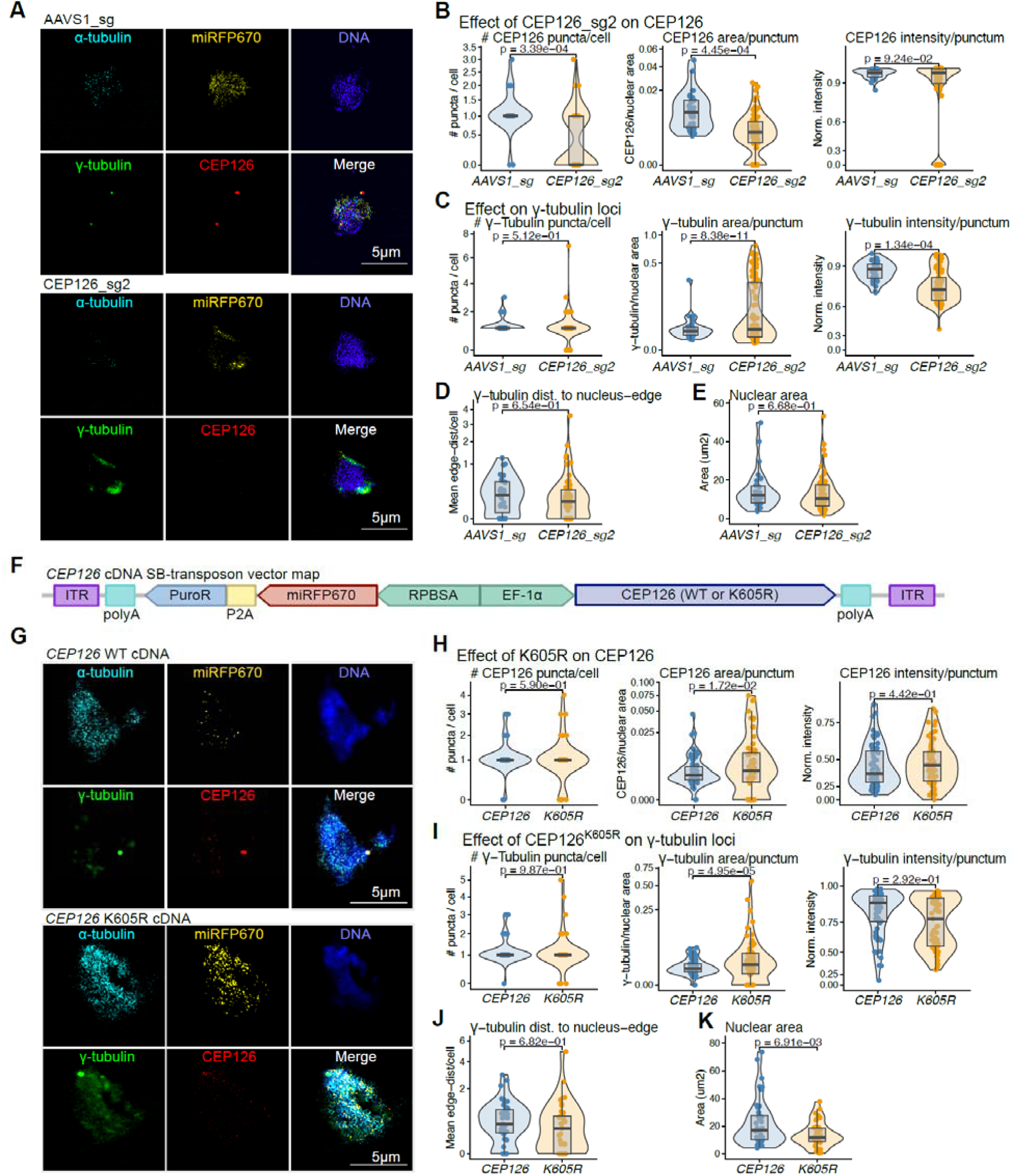
CEP126 loss and the familial CEP126 K605R variant alter centrosome-associated structures in neural progenitor cells. **(A)** Representative immunofluorescence images of AAVS1-targeting control NPCs and *CEP126*-sgRNA NPCs stained for α-tubulin, γ-tubulin, CEP126, miRFP670, and DNA. Scale bars, 5 μm. **(B)** Quantification of CEP126-positive foci number per cell, CEP126 focal area normalized to nuclear area, and CEP126 focal intensity in AAVS1-sgRNA and *CEP126*-sgRNA NPCs. **(C)** Quantification of γ-tubulin-positive foci number per cell, γ-tubulin focal area normalized to nuclear area, and γ-tubulin focal intensity in AAVS1-sgRNA and *CEP126*-sgRNA NPCs. **(D)** Quantification of the mean distance of γ-tubulin foci to the nuclear edge. **(E)** Quantification of nuclear area. **(F)** Schematic of the Sleeping Beauty transposon vector used to express WT *CEP126*or the familial *CEP126* K605R variant together with miRFP670 and puromycin resistance. **(G)** Representative immunofluorescence images of NPCs expressing WT *CEP126*cDNA or *CEP126* K605R cDNA stained for α-tubulin, γ-tubulin, CEP126, miRFP670, and DNA. Scale bars, 5 μm. **(H)**Quantification of CEP126-positive foci number per cell, CEP126 focal area normalized to nuclear area, and CEP126 focal intensity in WT *CEP126*- and *CEP126* K605R-expressing NPCs. **(I)** Quantification of γ-tubulin-positive foci number per cell, γ-tubulin focal area normalized to nuclear area, and γ-tubulin focal intensity in WT *CEP126*- and *CEP126* K605R-expressing NPCs. **(J)** Quantification of the mean distance of γ- tubulin foci to the nuclear edge. **(K)** Quantification of nuclear area. Violin plots show cell-level measurements with embedded boxplots. Statistical comparisons were performed using generalized linear mixed-effects models with condition as a fixed effect and biological replicate as a random intercept. Foci counts were modeled using negative-binomial regression with a log link, positive continuous area and distance measurements were modeled using Gamma regression with a log link, and normalized intensity measurements bounded between 0 and 1 were modeled using beta regression with a logit link. P values represent two-sided model-based contrasts between the indicated groups.

Quantitative image analysis revealed that *CEP126* disruption was associated with significant changes in centrosome- and microtubule-related cellular features. In *CEP126*-sgRNA cells, the γ-tubulin puncta had significantly greater area, but had decreased focal mean intensity measurements relative to control cells (**Figure 4C**). These changes occurred without affecting the number of puncta per cell or the distance of the γ-tubulin puncta to the nuclear edge. Additionally, there were no significant changes to nuclear area (**Figure 4D-E**).

Together, these data validate CEP126 depletion in edited NPCs and identify accompanying alterations in centrosome/cytoskeletal organization. These findings are consistent with the known role of CEP126 in γ-tubulin recruitment and suggest that the growth advantage observed after CEP126 disruption is accompanied by measurable changes in cellular architecture.

### The familial *CEP126* K605R variant disrupts centrosome-associated focal architecture

To determine whether the familial *CEP126* K605R variant was sufficient to alter these centrosome-associated phenotypes, we introduced either WT *CEP126* or *CEP126* K605R cDNA into NPCs using a Sleeping Beauty transposon vector co-expressing miRFP670 and a puromycin-resistance cassette (**Figure 4F**). NPCs expressing WT or K605R *CEP126* were identified by miRFP670 signal and analyzed using the same immunofluorescence-based workflow used above (**Figure 4G**). In contrast to *CEP126*-sgRNA cells, K605R-expressing cells did not show a reduction in *CEP126* foci number or mean *CEP126* focal intensity relative to WT *CEP126*-expressing cells. However, *CEP126* K605R expression significantly increased the area of CEP126-positive foci, indicating that the variant altered the morphology or organization of CEP126-associated structures without causing global loss of CEP126 signal (**Figure 4H**).

We next asked whether the K605R variant altered γ-tubulin-positive centrosomal structures. *CEP126* K605R did not significantly change the number of γ-tubulin foci per cell or the mean γ- tubulin focal intensity, but it significantly increased γ-tubulin focal area (**Figure 4I**). The distance of γ-tubulin foci to the nuclear edge was unchanged, suggesting that the variant affected focal morphology rather than gross centrosome positioning (**Figure 4J**). Nuclear area was also significantly reduced in K605R-expressing cells, indicating an additional effect on nuclear or cell-state-associated morphology (**Figure 4K**). Together, these cDNA-expression experiments show that *CEP126* K605R is sufficient to reproduce a subset of the centrosome-associated phenotypes observed after *CEP126* depletion, most notably enlargement of CEP126- and γ- tubulin-associated foci, while leaving foci number and centrosome-nuclear positioning largely unchanged. Thus, CEP126 loss and the familial K605R variant converge on altered centrosome- associated focal architecture, supporting the interpretation that K605R may produce partial functional impairment rather than complete loss of CEP126 expression.

## DISCUSSION

The genetic basis of familial GBM remains difficult to resolve because most affected kindreds are not explained by established cancer-predisposition syndromes or common glioma risk alleles. Although rare inherited variants can be identified by sequencing, distinguishing incidental private variation from biologically meaningful susceptibility genes requires functional evidence. In this study, we investigated a family with multiple GBM cases for suspected shared germline susceptibility. By integrating whole-genome sequencing, rare-variant filtering, and pooled CRISPR-Cas9 screening in human neural progenitor cells, we identified *CEP126* as the most robust candidate gene whose disruption conferred a reproducible selective advantage in vitro and in vivo.

CEP126 is a centrosomal protein previously implicated in primary cilium formation, but its role in cancer has been largely undefined. The identification of *CEP126* in this study suggests that centrosome- and cilia-associated biology may represent an underappreciated axis of inherited GBM susceptibility. Functional validation further supports a role for *CEP126* in regulating NPC fitness. *CEP126* disruption altered proliferation dynamics, but the phenotype was not simply an increase in the fraction of actively cycling cells. Rather, *CEP126* loss was associated with decreased apoptosis and enhanced tumorigenic growth in xenograft models. This suggests a model in which impaired *CEP126* function promotes NPC persistence and survival, potentially allowing cells to withstand stress or remain competent for subsequent oncogenic transformation. Such a mechanism would be consistent with inherited predisposition, in which a germline variant may create a permissive state that increases the probability of malignant progression after additional somatic events.

The imaging data provide a plausible cellular mechanism linking *CEP126* perturbation to altered progenitor-cell behavior. *CEP126* depletion reduced CEP126 protein signal and changed γ- tubulin-associated centrosomal features, including puncta area and intensity. Complementary cDNA-expression experiments showed that the familial K605R variant did not reduce CEP126 abundance, but reproduced a subset of the structural phenotype, including increased CEP126- and γ-tubulin-associated focal area. Because γ-tubulin is central to centrosome function, microtubule nucleation, spindle organization, and chromosome segregation, these findings raise the possibility that CEP126 disruption alters centrosome-associated architecture in ways that affect NPC survival or mitotic fitness. The K605R data make the important connection from the inherited missense variant to a measurable cellular phenotype, supporting partial functional alteration rather than simple loss of expression.

Several limitations should be considered. First, this is a single-family discovery study, and larger familial glioma cohorts will be required to determine whether *CEP126* variants are enriched among affected kindreds. Second, because matched normal DNA was unavailable for two affected relatives, germline variants in those individuals were inferred from tumor sequencing, although the analysis was restricted to shared variants consistent with constitutional inheritance. Third, the CRISPR experiments modeled *CEP126* loss of function rather than the endogenous K605R missense variant. Although the cDNA data support a functional effect of K605R, overexpression does not reproduce native dosage or regulation at the endogenous locus. Finally, the heterotopic xenograft model establishes tumorigenic potential but does not recapitulate the orthotopic brain microenvironment.

Future work should directly model the *CEP126*^K605R^ variant using knock-in and rescue systems, compare wild-type and variant CEP126 localization and function at endogenous expression levels, and define downstream effects on ciliation, centrosome organization, mitotic fidelity, and genomic stability. Orthotopic models and larger genetic datasets will be essential to establish disease relevance. Nonetheless, these findings nominate *CEP126* as a biologically tractable candidate GBM predisposition gene and demonstrate the value of functional screening as a bridge between rare familial variation and mechanistic cancer biology.

## CONCLUSION

This study identifies *CEP126* as a candidate familial GBM predisposition gene through integrated germline-variant discovery and functional CRISPR screening in human NPCs. *CEP126* disruption enhanced NPC survival, promoted tumorigenic growth, and altered centrosome-associated architecture, while CEP126 K605R reproduced part of this structural phenotype. These findings provide an experimental strategy for prioritizing rare inherited cancer-susceptibility variants and support future genetic and mechanistic studies of *CEP126* in familial gliomagenesis.

## Acknowledgements

P.R. was supported by the Cancer Biology Training Program Fellowship of the Yale Cancer Center (T32GM007205). S.C. is supported by Cancer Research Institute Lloyd J. Old STAR Award (CRI4964, CRI13945), Alliance for Cancer Gene Therapy (ACGT), NIH/NCI (R33CA281702), DoD (W81XWH-21-1-0514, HT9425-23-1-0472, HT9425-23-1-0860), Pershing Square Sohn Cancer Research Alliance, Prostate Cancer Foundation, and YCC Team Science Award. RGWV received support from NIH grants R21-CA256575, R01-CA271601; a grant from the Korea-US Collaborative Research Fund (KUCRF), funded by the Ministry of Science and ICT and Ministry of Health & Welfare, Republic of Korea (grant number: RS-2024- 00468725); a grant from the Lindonlight Collective. We also thank the Yale Center for Genomic Analysis for next generation sequencing; the W.M. Keck Foundation Biotechnology Resource Laboratory at Yale University for synthesizing the primers and oligonucleotides used in this study; and the Keck DNA sequencing facility at Yale for their assistance with Sanger DNA sequencing. Research reported in this publication was supported by the National Cancer Institute and the National Institute of General Medical Sciences of the National Institutes of Health under award numbers P30CA016359 and 1S10OD030363-01A1. Experiment schematics were created using BioRender.

## DATA AVAILABILITY

Source data and statistics for non-high-throughput experiments are provided in Supplemental Tables. Raw sequencing data for the screens will be deposited to the Gene Expression Omnibus (GEO) with pending accession number(s), and original code will be deposited to GitHub (https://github.com/Prenauer). Other relevant information or data are available from the corresponding author upon reasonable request.

## REFERENCES

1 Price, M. et al. CBTRUS Statistical Report: Primary Brain and Other Central Nervous System Tumors Diagnosed in the United States in 2018–2022. Neuro-Oncology 27, iv1–iv66 (2025). 10.1093/neuonc/noaf194

2 Weller, M., et al. Glioma. Nature Reviews Disease Primers 10, 33 (2024). 10.1038/s41572-024-00516-y

3 Wen, P. Y. et al. Glioblastoma in adults: A Society for Neuro-Oncology (SNO) and European Society of Neuro-Oncology (EANO) consensus review on current management and future directions. Neuro-Oncology 27, 2751–2788 (2025). 10.1093/neuonc/noaf177

4 Goldgar, D. E., Easton, D. F., Cannon-Albright, L. A. & Skolnick, M. H. Systematic population-based assessment of cancer risk in first-degree relatives of cancer probands. J Natl Cancer Inst 86, 1600–1608 (1994). 10.1093/jnci/86.21.1600

5 Malmer, B. et al. Genetic epidemiology of glioma. Br J Cancer 84, 429–434 (2001). 10.1054/bjoc.2000.1612

6 Malmer, B. et al. GLIOGENE an International Consortium to Understand Familial Glioma. Cancer Epidemiol Biomarkers Prev 16, 1730–1734 (2007). 10.1158/1055-9965.Epi-07-0081

7 Ostrom, Q. T. et al. The epidemiology of glioma in adults: a “state of the science” review. Neuro Oncol 16, 896–913 (2014). 10.1093/neuonc/nou087

8 Vijapura, C. et al. Genetic Syndromes Associated with Central Nervous System Tumors. Radiographics 37, 258–280 (2017). 10.1148/rg.2017160057

9 Melin, B. S. et al. Genome-wide association study of glioma subtypes identifies specific differences in genetic susceptibility to glioblastoma and non-glioblastoma tumors. Nat Genet 49, 789–794 (2017). 10.1038/ng.3823

10 Goodenberger, M. L. & Jenkins, R. B. Genetics of adult glioma. Cancer Genet 205, 613–621 (2012). 10.1016/j.cancergen.2012.10.009

11 Shete, S. et al. Genome-wide association study identifies five susceptibility loci for glioma. Nat Genet 41, 899–904 (2009). 10.1038/ng.407

12 Wrensch, M. et al. Variants in the CDKN2B and RTEL1 regions are associated with high-grade glioma susceptibility. Nat Genet 41, 905–908 (2009). 10.1038/ng.408

13 Eckel-Passow, J. E. et al. Inherited genetics of adult diffuse glioma and polygenic risk scores—a review. Neuro-Oncology Practice 9, 259–270 (2022). 10.1093/nop/npac017

14 Choi, D. J. et al. The genomic landscape of familial glioma. Sci Adv 9, eade2675 (2023). 10.1126/sciadv.ade2675

15 Bainbridge, M. N. et al. Germline mutations in shelterin complex genes are associated with familial glioma. J Natl Cancer Inst 107, 384 (2015). 10.1093/jnci/dju384

16 Jalali, A. et al. Targeted sequencing in chromosome 17q linkage region identifies familial glioma candidates in the Gliogene Consortium. Sci Rep 5, 8278 (2015). 10.1038/srep08278

17 Behan, F. M. et al. Prioritization of cancer therapeutic targets using CRISPR–Cas9 screens. Nature 568, 511–516 (2019). 10.1038/s41586-019-1103-9

18 Kellman, L. N. et al. Functional analysis of cancer-associated germline risk variants. Nature Genetics 57, 718–728 (2025). 10.1038/s41588-024-02070-5

19 Law, P. J. et al. Systematic prioritization of functional variants and effector genes underlying colorectal cancer risk. Nature Genetics 56, 2104–2111 (2024). 10.1038/s41588-024-01900-w

20 Chow, L. M., et al. Cooperativity within and among Pten, p53, and Rb pathways induces high- grade astrocytoma in adult brain. Cancer Cell 19, 305–316 (2011). 10.1016/j.ccr.2011.01.039

21 Friedmann-Morvinski, D. et al. Dedifferentiation of neurons and astrocytes by oncogenes can induce gliomas in mice. Science 338, 1080–1084 (2012). 10.1126/science.1226929

22 Marumoto, T. et al. Development of a novel mouse glioma model using lentiviral vectors. Nature Medicine 15, 110–116 (2009). 10.1038/nm.1863

23 Holland, E. C. Gliomagenesis: genetic alterations and mouse models. Nat Rev Genet 2, 120–129 (2001). 10.1038/35052535

24 Van Der Auwera, G. A. et al. From FastQ Data to High-Confidence Variant Calls: The Genome Analysis Toolkit Best Practices Pipeline. Current Protocols in Bioinformatics 43 (2013). 10.1002/0471250953.bi1110s43

25 Cibulskis, K. et al. Sensitive detection of somatic point mutations in impure and heterogeneous cancer samples. Nat Biotechnol 31, 213–219 (2013). 10.1038/nbt.2514

26 Richards, S. et al. Standards and guidelines for the interpretation of sequence variants: a joint consensus recommendation of the American College of Medical Genetics and Genomics and the Association for Molecular Pathology. Genetics in Medicine 17, 405–423 (2015). 10.1038/gim.2015.30

27 Melin, B. S. et al. Genome-wide association study of glioma subtypes identifies specific differences in genetic susceptibility to glioblastoma and non-glioblastoma tumors. Nature Genetics 49, 789–794 (2017). 10.1038/ng.3823

28 Mclaren, W. et al. The Ensembl Variant Effect Predictor. Genome Biology 17 (2016). 10.1186/s13059-016-0974-4

29 Auton, A. et al. A global reference for human genetic variation. Nature 526, 68–74 (2015). 10.1038/nature15393

30 Nomura, M. et al. The multilayered transcriptional architecture of glioblastoma ecosystems. Nature Genetics 57, 1155–1167 (2025). 10.1038/s41588-025-02167-5

31 Spitzer, A. et al. Deciphering the longitudinal trajectories of glioblastoma ecosystems by integrative single-cell genomics. Nature Genetics 57, 1168–1178 (2025). 10.1038/s41588-025-02168-4

32 Cheng, J. et al. Accurate proteome-wide missense variant effect prediction with AlphaMissense. Science 381, eadg7492 (2023). 10.1126/science.adg7492

33 Avsec, Ž., et al. Advancing regulatory variant effect prediction with AlphaGenome. Nature 649, 1206–1218 (2026). 10.1038/s41586-025-10014-0

34 Cancer Genome Atlas Research, N. Comprehensive genomic characterization defines human glioblastoma genes and core pathways. Nature 455, 1061–1068 (2008). 10.1038/nature07385

35 Li, M. M. et al. Points to consider for reporting of germline variation in patients undergoing tumor testing: a statement of the American College of Medical Genetics and Genomics (ACMG). Genetics in Medicine 22, 1142–1148 (2020). 10.1038/s41436-020-0783-8

36 Linehan, W. M. et al. Comprehensive Molecular Characterization of Papillary Renal-Cell Carcinoma. N Engl J Med 374, 135–145 (2016). 10.1056/NEJMoa1505917

37 Miller, K. D., et al. Brain and other central nervous system tumor statistics, 2021. CA: A Cancer Journal for Clinicians 71, 381–406 (2021). 10.3322/caac.21693

38 Karczewski, K. J. et al. The mutational constraint spectrum quantified from variation in 141,456 humans. Nature 581, 434–443 (2020). 10.1038/s41586-020-2308-7

39 Taipale, M. Disruption of protein function by pathogenic mutations: common and uncommon mechanisms. Biochem Cell Biol 97, 46–57 (2019). 10.1139/bcb-2018-0007

40 Churchhouse MD, A. et al. P076 Dock2 acts as a tumour suppressor via immune cell regulation of IDO1 in Inflammatory Bowel Disease–associated colorectal cancer. Journal of Crohn’s and Colitis 18, i343–i343 (2024). 10.1093/ecco-jcc/jjad212.0206

41 Schneider, T. et al. Large HERCs Function as Tumor Suppressors. Frontiers in Oncology Volume 9–2019 (2019). 10.3389/fonc.2019.00524

42 Burnichon, N. et al. MAX mutations cause hereditary and sporadic pheochromocytoma and paraganglioma. Clin Cancer Res 18, 2828–2837 (2012). 10.1158/1078-0432.Ccr-12-0160

43 Augert, A. et al. MAX Functions as a Tumor Suppressor and Rewires Metabolism in Small Cell Lung Cancer. Cancer Cell 38, 97–114.e117 (2020). 10.1016/j.ccell.2020.04.016

44 Davis, T. B. et al. PTPRS Regulates Colorectal Cancer RAS Pathway Activity by Inactivating Erk and Preventing Its Nuclear Translocation. Scientific Reports 8, 9296 (2018). 10.1038/s41598-018-27584-x

45 Briones-Orta, M. A. et al. Arkadia Regulates Tumor Metastasis by Modulation of the TGF-β Pathway. Cancer Research 73, 1800–1810 (2013). 10.1158/0008-5472.Can-12-1916

46 Wen, Z.-p., et al. Knockdown ATG4C inhibits gliomas progression and promotes temozolomide chemosensitivity by suppressing autophagic flux. Journal of Experimental & Clinical Cancer Research 38, 298 (2019). 10.1186/s13046-019-1287-8

47 Zhu, W. et al. WNK1-OSR1 kinase-mediated phospho-activation of Na+-K+-2Cl- cotransporter facilitates glioma migration. Mol Cancer 13, 31 (2014). 10.1186/1476-4598-13-31

48 Rahman, N. Realizing the promise of cancer predisposition genes. Nature 505, 302–308 (2014). 10.1038/nature12981

49 Koga, T. et al. Longitudinal assessment of tumor development using cancer avatars derived from genetically engineered pluripotent stem cells. Nature Communications 11 (2020). 10.1038/s41467-020-14312-1

50 Wang, X. et al. Sequential fate-switches in stem-like cells drive the tumorigenic trajectory from human neural stem cells to malignant glioma. Cell Research 31, 684–702 (2021). 10.1038/s41422-020-00451-z

51 Yang, L. et al. OR7A10 GPCR engineering boosts CAR-NK therapy against solid tumours. Nature 652, 740–751 (2026). 10.1038/s41586-026-10149-8

52 Bonavita, R. et al. Cep126 is required for pericentriolar satellite localisation to the centrosome and for primary cilium formation. Biology of the Cell 106, 254–267 (2014). 10.1111/boc.201300087

53 Johnson, K. C. et al. Acquired genetic and cell-state changes in IDH-mutant glioma progression. Nature (2026). 10.1038/s41586-026-10612-6

54 Zeng, K. et al. SEC61G assists EGFR-amplified glioblastoma to evade immune elimination. Proc Natl Acad Sci U S A 120, e2303400120 (2023). 10.1073/pnas.2303400120

55 Shi, Y. et al. Tumour-associated macrophages secrete pleiotrophin to promote PTPRZ1 signalling in glioblastoma stem cells for tumour growth. Nat Commun 8, 15080 (2017). 10.1038/ncomms15080

56 Plate, K. H., Breier, G., Weich, H. A. & Risau, W. Vascular endothelial growth factor is a potential tumour angiogenesis factor in human gliomas in vivo. Nature 359, 845–848 (1992). 10.1038/359845a0

57 Chen, S. et al. A genomic mutational constraint map using variation in 76,156 human genomes. Nature 625, 92–100 (2024). 10.1038/s41586-023-06045-0

58 Bonavita, R. et al. Cep126 is required for pericentriolar satellite localisation to the centrosome and for primary cilium formation. Biol Cell 106, 254–267 (2014). 10.1111/boc.201300087

59 Hendrickson, T. W., Yao, J., Bhadury, S., Corbett, A. H. & Joshi, H. C. Conditional mutations in gamma-tubulin reveal its involvement in chromosome segregation and cytokinesis. Mol Biol Cell 12, 2469–2481 (2001). 10.1091/mbc.12.8.2469

